# 3-STEP MODEL- AN EXPLORATIVE NOVEL APPROACH TO CLASSIFY SEPSIS: A LONGITUDINAL OBSERVATIONAL STUDY

**DOI:** 10.1101/2024.08.07.24311597

**Authors:** Jaideep Pilania, Prasan Kumar Panda, Ananya Das, Udit Chauhan, Ravikant

## Abstract

**Introduction:** Sepsis remains a critical healthcare challenge worldwide, demanding prompt identification and treatment to improve patient outcomes. Given the absence of a definitive gold standard diagnostic test, there is an imperative need for adjunct diagnostic tools to aid in early sepsis detection and guide effective treatment strategies. This study introduces a novel 3-step model to identify and classify sepsis, integrating current knowledge and clinical guidelines to enhance diagnostic precision.

**Methods:** This longitudinal observational study was conducted at a tertiary care teaching hospital in northern India. Adult patients admitted with suspected sepsis underwent screening using predefined criteria. The 3-step model consisted of Step 1, assessing dysregulated host response using a National Early Warning Score-2 (NEWS-2) score of ≥6; Step 2, evaluating risk factors for infection; and Step 3, confirming infection presence through clinical, supportive, or confirmatory evidence. Patients were categorized into Asepsis, Possible sepsis, Probable sepsis, or Confirmed sepsis at various intervals during hospitalization.

**Results:** A total of 230 patients were included. Initial categorization on Day 1 showed 13.0% in Asepsis, 35.2% in Possible sepsis, 51.3% in Probable sepsis, and 0.4% in confirmed sepsis. By Day 7, shifts were observed with 49.7% in Asepsis, 9.5% in Possible sepsis, 25.4% in Probable sepsis, and 15.4% in confirmed sepsis. At discharge or death, categories were 60.4% Asepsis, 5.2% Possible sepsis, 21.7% Probable sepsis, and 12.6% Confirmed sepsis. Transitions between categories were noted throughout hospitalisation, demonstrating the dynamic nature of sepsis progression and response to treatment.

**Conclusion:** The 3-step model effectively stratifies sepsis status over hospitalization, facilitating early identification and classification of septic patients. This approach holds promise for enhancing diagnostic accuracy, guiding clinical decision-making, and optimizing antibiotic stewardship practices. Further validation across diverse patient cohorts and healthcare settings is essential to confirm its utility and generalizability.

## INTRODUCTION

### Background/ Rationale

Sepsis is a long-known clinical entity whose definition, diagnosis, management and prognosis have evolved considerably over the time. Historically, sepsis was first used by the physician Hippocrates, around 2700 years ago. It was derived from the Greek word “sepo,” meaning “I rot”, to describe what was believed to be an internal decay-process that happened to unlucky individuals [1]. The term, “sepsis” has been used widely for many decades; and hence, it has been associated with various definitions, and the term had been vaguely applied to many clinical syndromes.

To improve the ability to study sepsis, and to have a universally accepted definition, a convention of experts met in the year 1992 and formalized the definition of the term, sepsis [2]. During that time, “sepsis” was defined as “an inflammatory response to the infection”. The clinical diagnosis of sepsis was defined by the presence of >/=2 of the “Systemic Inflammatory Response Syndrome (SIRS)” criteria paired with a suspected or confirmed source of an infection [3]. SIRS criteria had 4 parameters-heart rate, respiratory rate/ pCO_2_, body temperature and neutrophils [4], each of which having a single point. The diagnosis of sepsis was determined by identifying a suspected infection along with clinical or microbiological evidence of same, accompanied by at least two out of the four criteria of systemic inflammatory response (SIRS) [5]. These proposed definitions were used for around a decade, but “The Sepsis-1” 1992 definitions faced several criticisms. One significant critique was that the SIRS criteria often represented a befitting bodily response to an infection rather than a pathological state, making it non-specific for patients with sepsis. The SIRS criteria emphasize upon the inflammatory response produced, which is a characteristic of numerous critical conditions such as trauma, pancreatitis, and postsurgical inflammation, which is not related to sepsis per se [6]. On application of the above criteria for sepsis, a large group of patients met criteria, and even more than 90% of the patients who gets admitted in an intensive care unit (ICU) facility met the sepsis criteria [7]. Additionally, the term “severe sepsis” was introduced to denote organ dysfunction resulting from sepsis [8].

In 2001, a second group of experts met to review and update the Sepsis-1 definitions [9]. Although the core definitions remained largely unchanged, they incorporated the “Sequential Organ Failure Assessment (SOFA)” criteria for better identification of organ dysfunction associated with severe sepsis. Sepsis was still defined as >/=2 SIRS criteria AND suspected infection, and severe sepsis, which was introduced as new term, was defined as sepsis AND organ dysfunction (change in SOFA >/=2 points). Hence, the initially proposed definition, as outlined in Sepsis-1 was widely utilized for nearly twenty years despite the criticism as highlighted above, due to non-availability of better definition or criteria for sepsis. Since inflammation is a normal and beneficial response to many infections, defining sepsis posed the challenge of distinguishing between the typical inflammatory response of a simple infection and the severe, dysregulated response that characterizes life-threatening sepsis [8].

In 2016, the Sepsis Task Force redefined sepsis as a life-threatening condition resulting from organ dysfunction caused by an abnormal host response to infection [10]. Clinically, this is identified by an acute increase of 2 or more points in the SOFA score when an infection is suspected [10,11]. With the updated definition, the term “severe sepsis” became redundant and was thus eliminated.

Over the time, various scoring systems were formulated which could predicts the sepsis related outcome in patients with sepsis such as “SOFA” (Sequential Organ Failure Assessment) score, “NEWS” (National Early Warning Score)/ “NEWS-2”, “qSOFA” (quick Sequential Organ Failure Assessment) score, “APACHE-II” (Acute Physiology and Chronic Health Evaluation-II) etc, and many more. All these scores mainly predict the severity of organ dysfunction produced by dysregulated host response; however, they can’t be directly used to diagnose sepsis. These scores are mainly used when an individual has suspected sepsis, with organ dysfunction to know or predict the prognosis or outcome, related to sepsis/ suspected sepsis. However, use of scoring systems has an increasing value for predicting the progression of patients to sepsis [12]. Apart from the various scoring systems, many biomarkers were also evaluated for their role in sepsis, diagnostic and therapeutic role.

Among various biomarkers, Procalcitonin and the C-reactive protein (CRP), are the two most widely studied biomarkers in various contexts of sepsis, including diagnosis and guiding antibiotics therapy. C-reactive protein (CRP) has been widely used by clinicians as a biomarker of inflammation for many decades. It is highly sensitive for sepsis, but it lacks specificity when compared with procalcitonin (PCT) [13].

With the passage of time, our understanding of the sepsis improved drastically in terms of its pathophysiology, organ dysfunction, clinical features and management with emerging evidences and ongoing many researches. This led to the formulation of latest sepsis definition and its management guidelines. As per the latest surviving sepsis campaign: International guidelines for management of sepsis and septic shock, 2021 (Sepsis-3), sepsis is defined as life-threatening organ dysfunction caused by a dysregulated host response to infection [10]. However, despite of our increased understanding, sepsis is associated with very high rates of mortality in both developed and developing countries even with the management. That’s why, The World Health Assembly and WHO made sepsis a global health priority in 2017, and have adopted a resolution to improve the prevention, diagnosis, and management of sepsis [14]. According to current data, there is no gold standard diagnostic test to diagnose sepsis.

Hence, sepsis remains a critical healthcare challenge worldwide, demanding prompt identification and treatment to improve patient outcomes. Given the absence of a definitive gold standard diagnostic test for sepsis, there is an imperative need for adjunct diagnostic tools to aid in early sepsis detection and guide effective treatment strategies.

### Objective of Study

With the objective of classification of sepsis into different categories on different intervals of hospitalisation this longitudinal observational study was done at a tertiary care teaching hospital in northern India. This study introduces a novel 3-step model to identify and classify sepsis, integrating current knowledge and clinical guidelines to enhance diagnostic precision. The novel 3-step method was utilised to classify patients into different sepsis categories (Asepsis, Possible sepsis, Probable sepsis and Confirm sepsis) at Day-1, Day-7, Day-14 and Day-28/ Discharge/ Death during the hospital stay. Apart from classification, changes among different sepsis categories with duration of hospital stay was also evaluated.

## METHODS

### Study Design

This was a longitudinal observational study

### Study Setting

It was done at a tertiary care teaching hospital in northern India in the department of general medicine from 1^st^ January to 31^st^ December, 2023, after the approval from Institute Ethics Committee (IEC). Data was collected from patients and was entered in RedCap software (AIIMS Rishikesh version), and also in the Microsoft Excel sheet.

### Objective

-To identify and classify sepsis in patients with suspected sepsis using a novel 3-step approach.

-Estimate changes in sepsis categories on different time intervals.

### Participants

Participants includes patients who were admitted in the department of general medicine with who were eligible as per inclusion and exclusion criteria of the study from 1^st^ January to 31^st^ December, 2023.

### Inclusion Criteria

**-**Patients of age >/= 18 years who are admitted in department of general medicine with suspected sepsis.

### Exclusion Criteria

-Patients who were diagnosed etiologically other than sepsis with 5 days of hospital admission

-Patients whose data was missing

### Variables/ Outcomes

-To estimate proportion of patients in different categories of sepsis at Day-1, Day-7, Day-14 and Day-28/ death/ discharge.

-To estimate change in category of sepsis with different days of observation.

### Data source

Patients admitted in general medicine ward, and also from medical records of discharged patients were used for data collection.

### Study size

The study size or sample size was not mathematically calculated, as no prior refence study was available for same. So, as per feasibility, universal sampling method was used for samples of the study.

### Statistical Method

The study was primarily an observational study which used descriptive data analysis. Patients with missing data were not included for the final result analysis.

## METHODOLOGY

-Patients admitted in Department of General Medicine with suspected sepsis were screened for inclusion and exclusion criteria of the study and patients fulfilling criteria were included in the study.

-Patients, both-admitted and discharged, were assessed for inclusion in the study. For discharged patients, data was extracted from available hospital records and then were subjected to 3 step approach/ model.

-A 3-step model was prepared using the sepsis definition, and were divided into various steps. Initial step included evidence of dysregulated host response, which was evaluated with the use of National Early Warning Score-2 (NEWS-2 score). As per the published report by Royal College of Physicians (RCP), a NEWS-2 score of 5 or more should make one think for sepsis [15]; thus, for our study, we took a higher value of NEWS-2 score as an evidence of dysregulated host response i.e a NEWS-2 score of ≥6 was used.

-“Suspected sepsis” term was used, which was defined by one the following parameters:

i. Need of antibiotics for management
ii. Evidence of infection anywhere in the body
iii. Organ dysfunction not explained by non-infective cause
iv. Patient improved after antibiotics.

The term of suspected sepsis, with any one of the above-mentioned points, was first validated by experts from various fields including internal medicine, infectious disease, etc, including both, from the institute experts and also not related to institute, and was then incorporated into the study.

-After screening and inclusion, patients were subjected to 3 step approach/ model and were followed, either physical or via available hospital data or records, till an outcome (which is discharge/ death) is reached, and were categorized into different sepsis categories on different day on follow-up.

-All baseline data was collected including vitals (for calculation of NEWS-2 score), laboratory data, cultures, and also the empirically used antibiotics.

-3-step approach/ model has step-1 as evidence of dysregulated host response (assessed by the use of NEWS-2 score), step-2 was risk factor of infection and final, step-3, was to look for evidence of infection.

### METHODOLOGY (cont.)

To identify and classify sepsis in a patient using novel 3-step model:

### STEP-1 EVIDENCE OF DYSREGULATED HOST RESPONSE

-NEWS-2 >/=6 was used as an evidence of dysregulated host response.

### STEP-2 RISK FACTOR FOR INFECTION

-Multiple risk factors are associated with sepsis, such as – decompensated chronic liver disease, dialysis dependent chronic kidney disease, uncontrolled diabetes mellitus, chronic pulmonary diseases, immunosuppressive states, older age, recent previous hospitalisation etc.

### STEP-3 EVIDENCE OF INFECTION: -

#### 3(A) CLINICAL EVIDENCE-SYNDROMIC DIAGNOSIS [16]

-Includes syndromic diagnosis such as pyelonephritis, infective endocarditis, meningitis, intra-abdominal infections, skin & soft tissue infections, pneumonia, osteomyelitis, etc

#### 3(B) SUPPORTIVE/ SUGGESTIVE EVIDENCE

**-(i) Imaging:** Showing evidence of infection like chest x-ray, ultrasound, computed tomography (CT Scan), magnetic resonance imaging (MRI) etc
**-(ii) Biomarkers:** Detected from samples such as blood (procalcitonin, beta-1,3-glucan, galactomannan), urine, other fluids etc.

#### 3(C) CONFIRMATORY EVIDENCE

**-(i) Direct Visualisation:** via Eye/Open Method i.e without use of any instrument like Myiasis, Ectoparasites etc.
**(ii) Endoscopic Evidence/ Visualisation**
**(iii) Microscopy & Culture Growth and Sensitivity-** Blood, Urine, Endotracheal Tube (ET) Aspirate/ Bronchoalveolar Lavage (BAL), Catheter Tip, Wound, Swab culture, Biopsy Material, Sputum, Other fluids like cerebrospinal fluid (CSF), Pleural, Pericardial, Ascitic, Synovial etc.
**(iv) Polymerase Chain Reaction (PCR)/ Gene Detection Methods**
**(v) Immunological Methods** like immunochromatography (ICT), Chemiluminescence immunoassay (CLIA), Enzyme-linked immunosorbent assay (ELISA) and others.

## 3-STEP MODEL FOR SEPSIS

**Figure-1:**
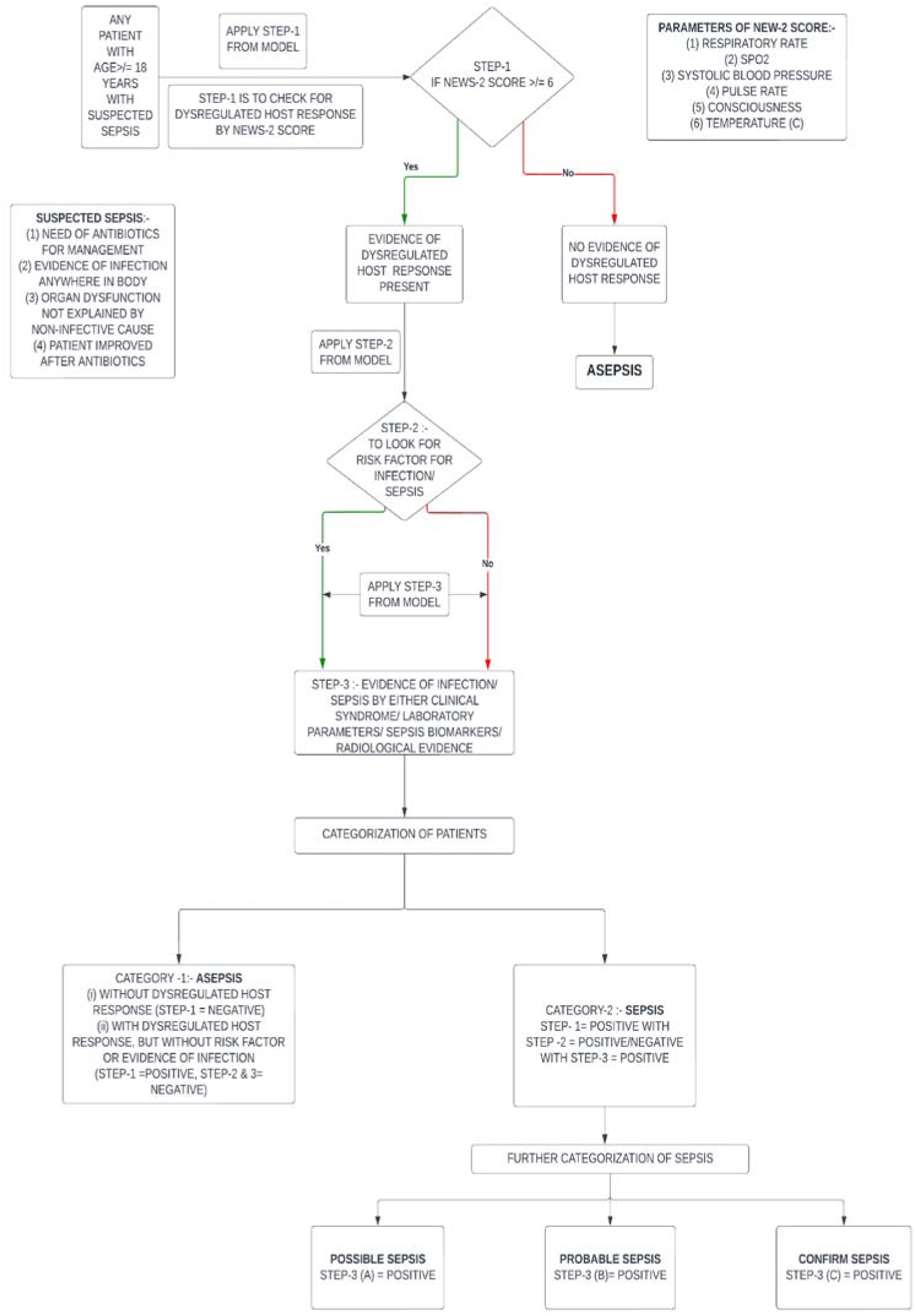
Flowchart to use the 3-Step model for sepsis classification.

## CATEGORIZATION OF SEPSIS

**Table-1:** Table showing various sepsis categories.

| **CATEGORY** | **INTERPRETATION** | **OUTCOME** |
| --- | --- | --- |
| CATEGORY-1 | (i) STEP-1 = NEGATIVE<br>(ii) STEP-1= POSITIVE WITH<br>STEP-2 & 3= NEGATIVE | **ASEPSIS** |
| CATEGORY-2 | STEP-1, 2 & 3(A) = POSITIVE | **POSSIBLE SEPSIS** |
| CATEGORY-3 | STEP-1, 2 & 3(B) = POSITIVE | **PROBABLE SEPSIS** |
| CATEGORY-4 | STEP-1, 2 & 3(C) = POSITIVE | **CONFIRM SEPSIS** |

## RESULTS

This was an observational longitudinal study done at tertiary teaching hospital in northern India which included patients with age >/= 18 years with suspected sepsis (inclusion criteria) who were admitted in the department of General Medicine. A total of 1867 patients were screened and after exclusion criteria, and patients with missing data were excluded, resulting in a study cohort of 230 patients for analysis.

The mean age (in years) was 40.70 ± 14.49 years, and out of the 230 participants, 113 (49.1%) were male and 117 (50.9%) were female, suggestive of slight female predominance. Age was further sub-divided into 3 main groups, with 113 (49.13%) patients belonging to 18-40 years of age group, 94 (40.87%) patients were in the 41-60 years of age group, and the remaining 23 (10%) patients were in the >60 years of age group, showing that majority of patients were from 18-40 years of age group [Table-2].

**Table-2:** Demographic Characteristics.

| Basic Details | Mean ± SD \|\| Median (IQR) \|\| Min-Max OR N (%) |
| --- | --- |
| Age (Years) | 40.70 ± 14.49 \|\| 41.00 (28.00-53.00) \|\| 18.00 - 70.00 |
| Age Group |  |
| 18-40 years | 113 (49.13%) |
| 41-60 years | 94 (40.87%) |
| >60 years | 23 (10%) |
| Gender |  |
| Male | 113 (49.1%) |
| Female | 117 (50.9%) |
| State |  |
| Uttar Pradesh | 120 (52.2%) |
| Uttarakhand | 98 (42.6%) |
| Bihar | 3 (1.3%) |
| Rajasthan | 2 (0.9%) |
| Delhi | 1 (0.4%) |
| Gujarat | 1 (0.4%) |
| Jharkhand | 1 (0.4%) |
| Madhya Pradesh | 1 (0.4%) |
| Punjab | 1 (0.4%) |
| Tamil Nadu | 1 (0.4%) |
| Telangana | 1 (0.4%) |

-Sepsis was categorised into different categories as per the novel 3-step model. Day-1 of classification was dominated by probable sepsis (51.3%) and possible sepsis (35.2%) which was later comprised mainly of asepsis (60.4%) category at the time of outcome. Table-3 to Table-6 show the proportion of patients in different categories of sepsis in Day-1, Day-7, Day-14 and Day-28/ discharge/ death (outcome). Table-7 shows the summary of sepsis categorization on different time intervals from day-1 to day of outcome.

**Table-3:**
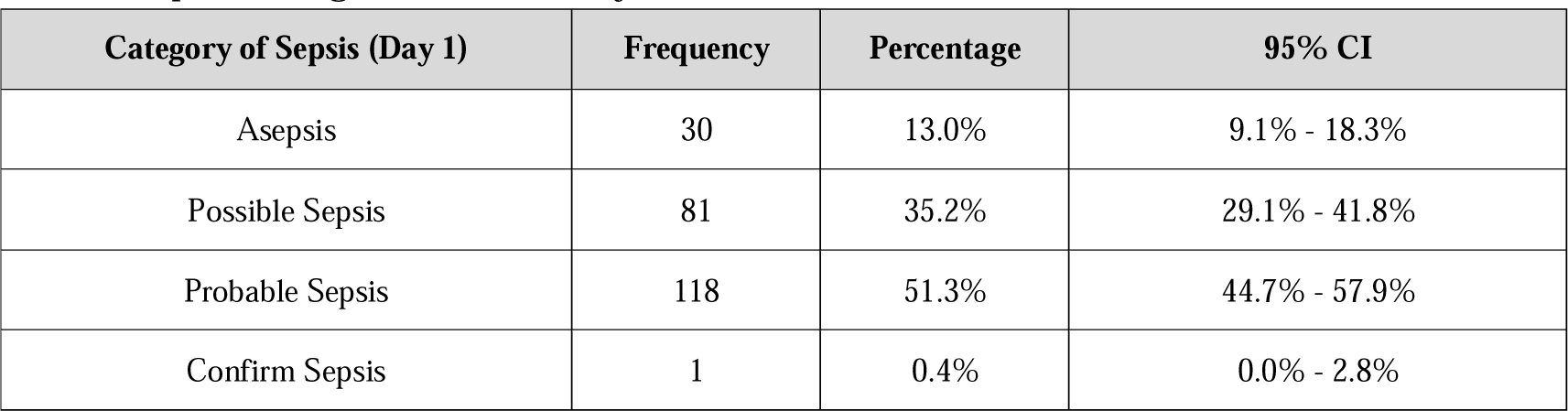
Sepsis Categorisation on Day-1.

**Table-4:** Sepsis Categorisation on Day-7.

| Category of Sepsis (Day 7) | Frequency | Percentage | 95% CI |
| --- | --- | --- | --- |
| Asepsis | 84 | 49.7% | 42.0% - 57.5% |
| Possible Sepsis | 16 | 9.5% | 5.7% - 15.2% |
| Probable Sepsis | 43 | 25.4% | 19.2% - 32.8% |
| Confirm Sepsis | 26 | 15.4% | 10.5% - 21.9% |

**Table-5:** Sepsis Categorisation on Day-14.

| Category of Sepsis (Day 14) | Frequency | Percentage | 95% CI |
| --- | --- | --- | --- |
| Asepsis | 40 | 54.8% | 42.8% - 66.3% |
| Possible Sepsis | 2 | 2.7% | 0.5% - 10.4% |
| Probable Sepsis | 21 | 28.8% | 19.1% - 40.7% |
| Confirm Sepsis | 10 | 13.7% | 7.1% - 24.2% |

**Table-6:** Sepsis Categorisation on Day of Outcome (Day-28/ Discharge/ Death)

| Category of Sepsis (At Outcome) | Frequency | Percentage | 95% CI |
| --- | --- | --- | --- |
| Asepsis | 139 | 60.4% | 53.8% - 66.7% |
| Possible Sepsis | 12 | 5.2% | 2.8% - 9.2% |
| Probable Sepsis | 50 | 21.7% | 16.7% - 27.7% |
| Confirm Sepsis | 29 | 12.6% | 8.7% - 17.8% |

**Table-7:** Summary of category of sepsis.

| Category of Sepsis | Asepsis | Possible Sepsis | Probable Sepsis | Confirm Sepsis |
| --- | --- | --- | --- | --- |
| **Day 1** | 30 (13.0%) | 81 (35.2%) | 118 (51.3%) | 1 (0.4%) |
| **Day 7** | 84 (49.7%) | 16 (9.5%) | 43 (25.4%) | 26 (15.4%) |
| **Day 14** | 40 (54.8%) | 2 (2.7%) | 21 (28.8%) | 10 (13.7%) |
| **At Outcome** | 139 (60.4%) | 12 (5.2%) | 50 (21.7%) | 29 (12.6%) |

-Apart from sepsis categorization into different categories, change from one category to different category was also evaluated with time course. It was observed that from day-1 to day-7, 18.9% and 23.1% patients belonging to probable and possible sepsis category respectively, moved to asepsis category. This trend was observed throughout the course of observation. Table-8 to Table-10 and Figure-2 to Figure-4 shows the change in sepsis category of patients at different time intervals w.r.t to day-1 category of sepsis.

**Table-8:** Change in sepsis category from Day-1 to Day-7.

| Category of Sepsis |  | Day 1 |  |  |  |  | Stuart-Maxwell test |  |
| --- | --- | --- | --- | --- | --- | --- | --- | --- |
| | | Asepsis | Possible Sepsis | Probable Sepsis | Confirm Sepsis | Total | $\chi^2$ | P Value |
| **Day 7** | **Asepsis** | 13 (7.7%) | 39 (23.1%) | 32 (18.9%) | 0 (0.0%) | 84 (49.7%) | 91.238 | <0.001 |
|  | **Possible Sepsis** | 0 (0.0%) | 6 (3.6%) | 10 (5.9%) | 0 (0.0%) | 16 (9.5%) |  |  |
|  | **Probable Sepsis** | 2 (1.2%) | 7 (4.1%) | 34 (20.1%) | 0 (0.0%) | 43 (25.4%) |  |  |
|  | **Confirm Sepsis** | 0 (0.0%) | 5 (3.0%) | 21 (12.4%) | 0 (0.0%) | 26 (15.4%) |  |  |
|  | **Total** | 15 (8.9%) | 57 (33.7%) | 97 (57.4%) | 0 (0.0%) | 169 (100.0%) |  |  |

The uncolored cells on the diagonal represent patients whose category did not change. The red shaded cells represent patients who moved to a lower category. The green shaded cells represent patients who moved to a higher category.

**Figure-2:**
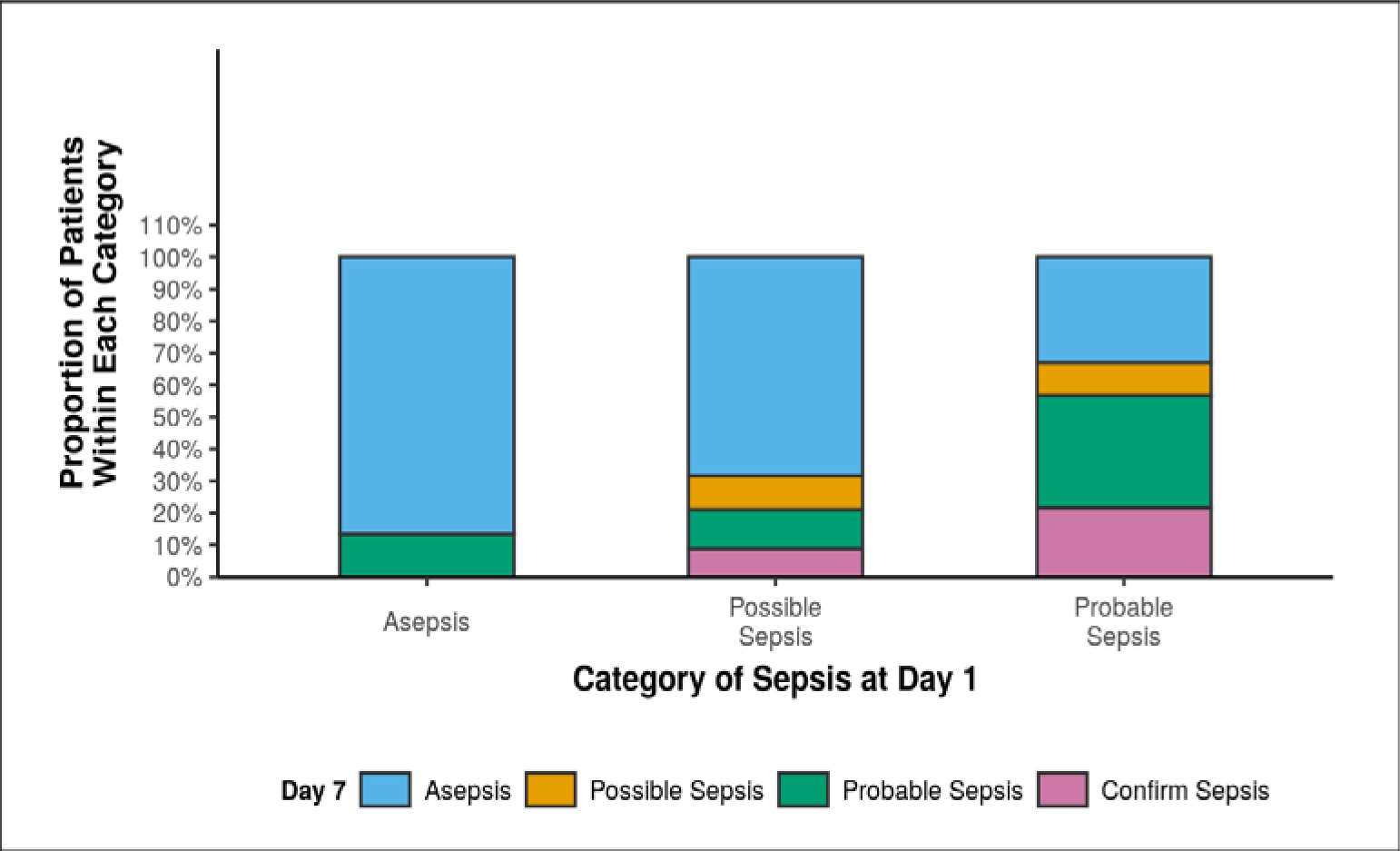
Graph showing change in sepsis category from Day-1 to Day-7.

**Table-9:** Change in sepsis category from Day-1 to Day-14.

| Category of Sepsis |  | Day 1 |  |  |  |  | Stuart-Maxwell test |  |
| --- | --- | --- | --- | --- | --- | --- | --- | --- |
| | | Asepsis | Possible Sepsis | Probable Sepsis | Confirm Sepsis | Total | $\chi^2$ | P Value |
| Day 14 | Asepsis | 2 (2.7%) | 17 (23.3%) | 21 (28.8%) | 0 (0.0%) | 40 (54.8%) | 45.153 | <0.001 |
|  | Possible Sepsis | 0 (0.0%) | 0 (0.0%) | 2 (2.7%) | 0 (0.0%) | 2 (2.7%) |  |  |
|  | Probable Sepsis | 1 (1.4%) | 2 (2.7%) | 18 (24.7%) | 0 (0.0%) | 21 (28.8%) |  |  |
|  | Confirm Sepsis | 0 (0.0%) | 2 (2.7%) | 8 (11.0%) | 0 (0.0%) | 10 (13.7%) |  |  |
|  | Total | 3 (4.1%) | 21 (28.8%) | 49 (67.1%) | 0 (0.0%) | 73 (100.0%) |  |  |

The uncolored cells on the diagonal represent patients whose category did not change. The red shaded cells represent patients who moved to a lower category. The green shaded cells represent patients who moved to a higher category.

**Figure-3:**
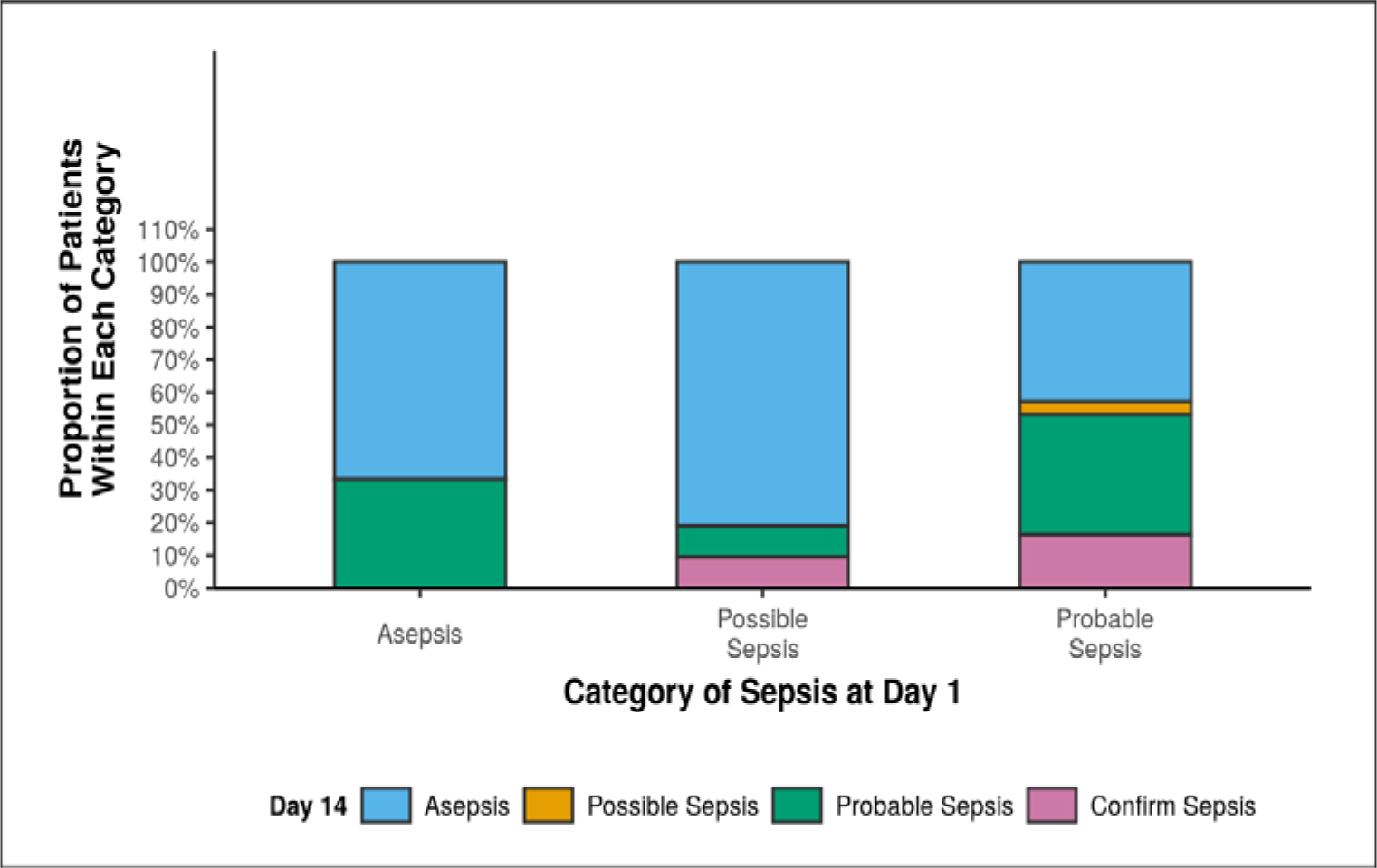
Graph showing change in sepsis category from Day-1 to Day-14.

**Table-10:** Change in sepsis category from Day-1 to Outcome.

| Category of Sepsis |  | Day 1 |  |  |  |  | Stuart-Maxwell test |  |
| --- | --- | --- | --- | --- | --- | --- | --- | --- |
| | | Asepsis | Possible Sepsis | Probable Sepsis | Confirm Sepsis | Total | $\chi^2$ | P Value |
| At Outcome | Asepsis | 29 (12.6%) | 66 (28.7%) | 44 (19.1%) | 0 (0.0%) | 139 (60.4%) | 135.117 | <0.001 |
|  | Possible Sepsis | 0 (0.0%) | 6 (2.6%) | 6 (2.6%) | 0 (0.0%) | 12 (5.2%) |  |  |
|  | Probable Sepsis | 1 (0.4%) | 3 (1.3%) | 46 (20.0%) | 0 (0.0%) | 50 (21.7%) |  |  |
|  | Confirm Sepsis | 0 (0.0%) | 6 (2.6%) | 22 (9.6%) | 1 (0.4%) | 29 (12.6%) |  |  |
|  | Total | 30 (13.0%) | 81 (35.2%) | 118 (51.3%) | 1 (0.4%) | 230 (100.0%) |  |  |

The uncolored cells on the diagonal represent patients whose category did not change. The red shaded cells represent patients who moved to a lower category. The green shaded cells represent patients who moved to a higher category.

**Figure-4:**
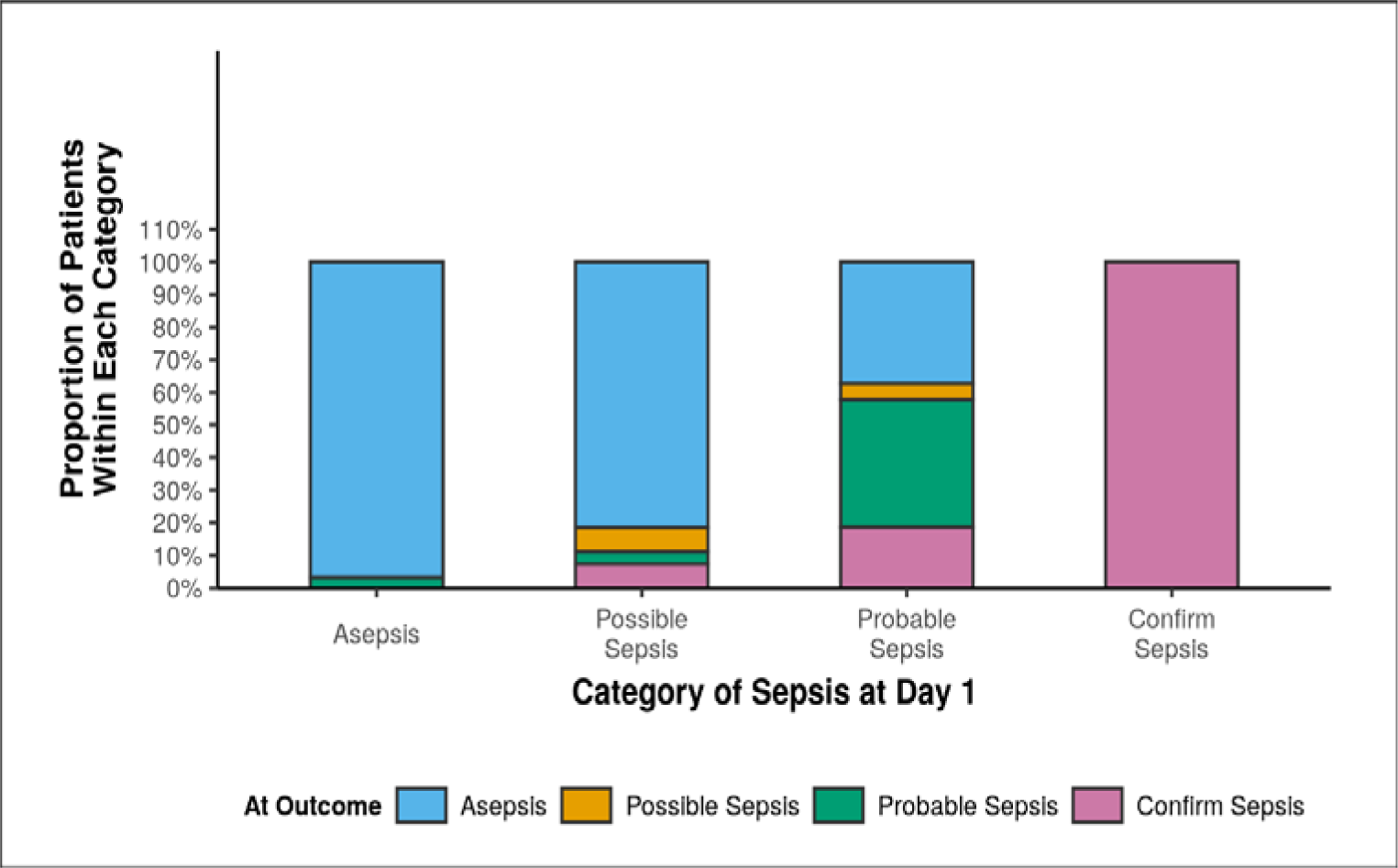
Graph showing change in sepsis category from Day-1 to Day-14.

Table-11 shows the summary of other parameters which were reported from study cohort, including days of hospitalization, NEWS-2 score, total ICU admissions, duration of ICU stay, outcome (discharge/ discharge with unstable vitals and death) and mortality.

Mean days of hospitalisation was 11.38 ± 7.58 days with mean NEWS-2 score being 4.08 ± 3.08. Among the study cohort of 230 patients, 23 (10.0%) were admitted in ICU with mean duration of ICU stay of 10.17 ± 13.21 days. The study had 2.1% of mortality rate with majority, which is 210 out of 230 or 91.3% patients were discharged with stable vitals.

**Table-11:**
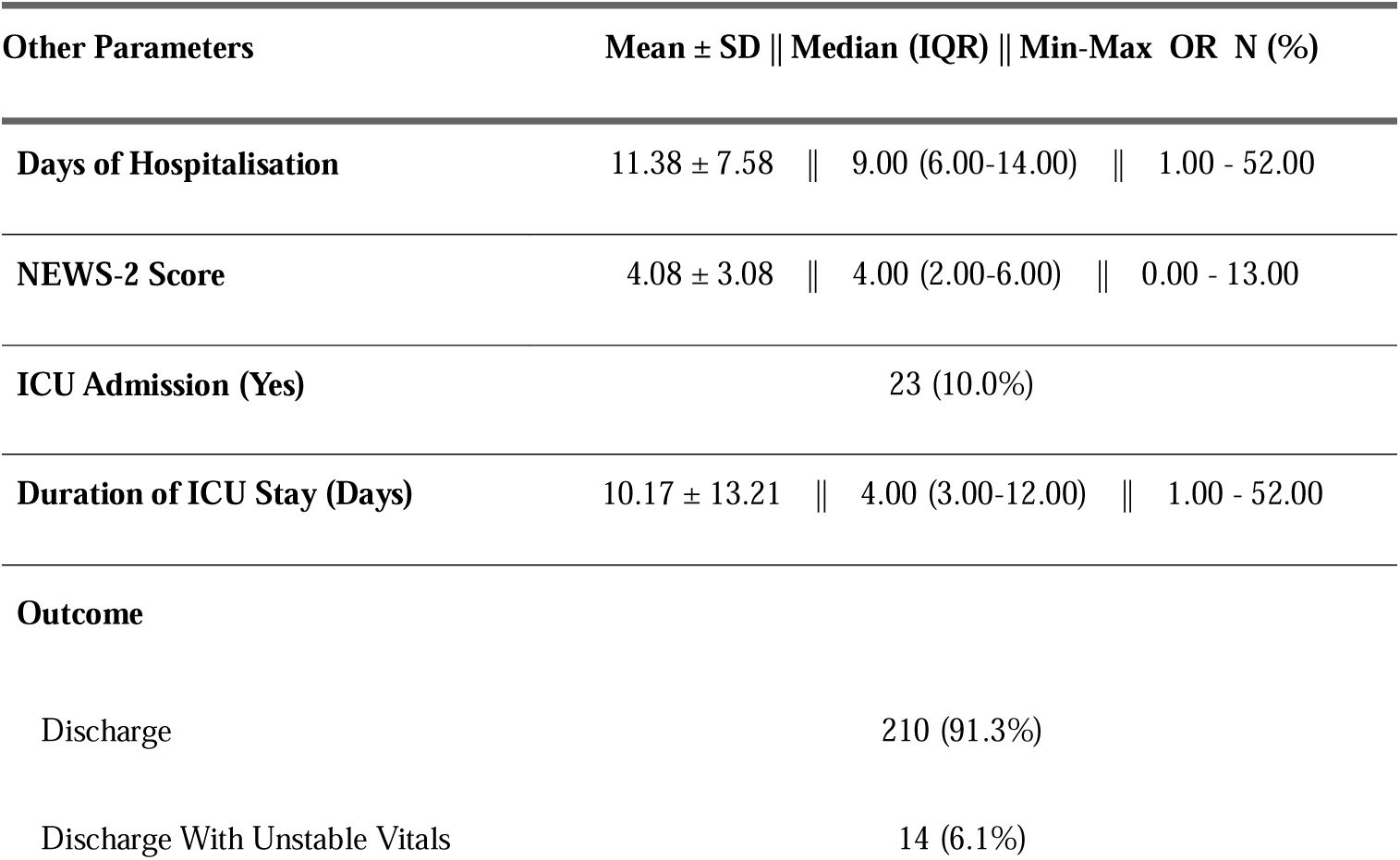
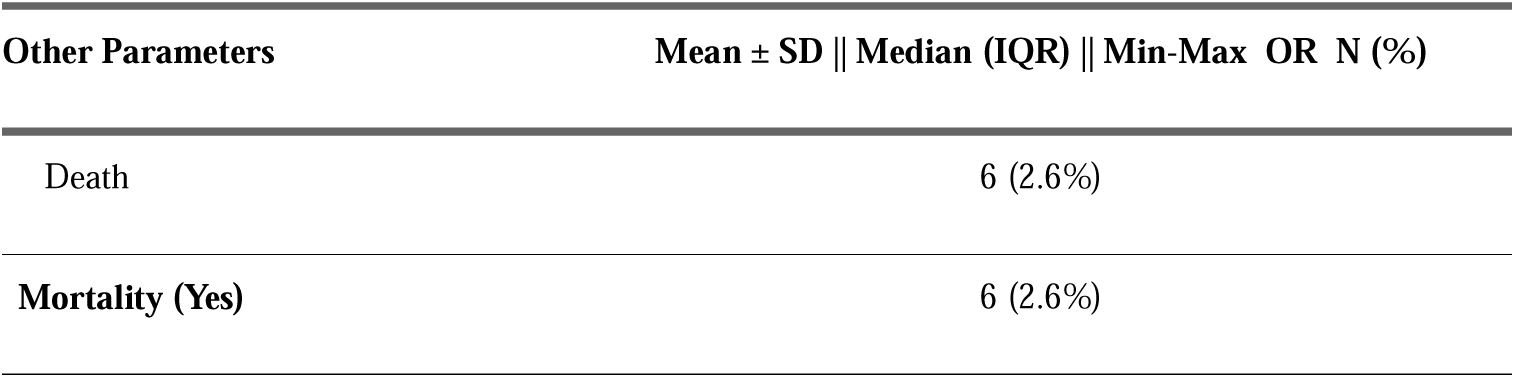
Summary of other parameters.

## DISCUSSION

This study was done with the aim to provide an aid to the clinicians in the diagnosis and classification of sepsis for better differentiation of patients with suspected sepsis and their prompt management. Sepsis has been made a global health priority in 2017 by WHO [14] due to the associated very high mortality rates with it, even after treatment. Understanding of sepsis has changed considerably over the last few decades with the advent of newer techniques to understand the molecular mechanism, aiding in the better understanding of the patho-mechanism of sepsis and associated organ dysfunction with it. This study was also aimed to contribute its tiny part in the better understanding and management of sepsis. Sepsis is defined as per the latest Surviving Sepsis guidelines of 2021 as life-threatening organ dysfunction caused by a dysregulated host response to infection [10].

This longitudinal observational study done at a tertiary center in Northern India including 230 patients of age >/= 18 years with suspected sepsis in Department of General Medicine used a novel 3-step approach for classification and identification of sepsis using NEWS-2 score. As per the updated report of working party published by Royal College Physicians (RCP) in December 2017 about executive summary and recommendations regarding National Early Warning Score-2 (NEWS-2) score mentioned that the use of NEWS-2 score of 5 or more should alert clinician about sepsis [17]. Being easy to use, NEWS-2 score was used in this study as it involves parameters which can be obtained with vitals of patients, negating use of any blood test or laboratory parameters, making it easy to calculate at bedside, even in resource limited setting.

The study had its background, rather its origin, from the lacuna in the definition of sepsis, given by latest guidelines, that sepsis is defined as life-threatening organ dysfunction caused by a dysregulated host response to infection, and to try to fill and explore this lacuna. Despite our understanding of sepsis, better therapeutic approach to manage sepsis; we still lack behind in diagnosing sepsis with certainty, as there is no gold standard test which can be used to diagnose sepsis. Hence, this 3 step-approach or model was created and used in this study for identification and classification/ categorization of sepsis into different categories. We used the two components of sepsis definition as two arms of our study, i.e dysregulated host response and evidence of infection, which was incorporated into the study using 3 step-model or approach.

This study is one of its kind, as no such study was done previously to use a stepwise approach for sepsis identification and classification, rather major chunk of studies are done using various available scoring systems for comparing the mortality, efficacy and prognostic accuracy of such scoring systems in sepsis.

The study included a total of 230 patients, with mean age of 40.7 ± 14.49 years with slight dominance of female gender, 117 (50.9%) females and 113 (49.1%) males, and majoring of patients were from Uttar Pradesh (UP) and Uttarakhand (UK) states with minority from other states [Table-2], with mean duration of hospital stay/ hospitalization of 11.38 ± 7.58 days.

The objective of the study was to classify the patients into various proposed categories of sepsis on the basis of 3 step approach/ model and also to assess the change in category of the patient with its course of hospitalization/ duration of hospital admission. It was observed that at the time of admission (day-1), on the basis of clinician’s judgement and scoring system, majority of patients were classified into category of probable sepsis (51.3%), followed by possible sepsis (35.2%) and asepsis (13.0%) category [Table-3]. This distribution can be justified or explained on the basis that at the time of initial evaluation by treating clinician, it is sometimes very difficult to accurately predict or identify patients with sepsis. It was observed that during the course of hospital stay, there was significant changes in proportion of patients from one category to another, i.e dynamic change of sepsis category, which was evaluated at different days of admission, such as at day of admission, at day-7, at day-14 and till outcome (death/ discharge) is reached.

By the day-7 of admission, the majority of patients were reclassified into different sepsis categories, now, majority were put into asepsis category (49.7%), followed by probable sepsis (25.4%), confirm sepsis (15.4%) and possible sepsis (9.5%). Similar trend was observed at day-14, with majority of patients again belonging to asepsis group (54.8%), followed by probable sepsis group (28.8%), confirm sepsis group (13.7%) and possible sepsis (2.7%) [Table-4 & 5 respectively].

This change in the sepsis categories over the course of time can be attributed to right diagnostic approach used by treating physician at the time of presentation, right use of empirical antibiotics for a right duration, which led to this dynamic change in the category of sepsis, from initial predominant category of probable and possible sepsis to final category in majority of patients being asepsis. Another explanation for this change of category can be attributed to the initial wrong categorization of sepsis, when patients were actually having organ dysfunction due to non-infectious cause, which was later identified over their course of hospital stay, or there can be single organ involvement only due to a non-infectious etiology, which prompted clinician/ treating physician to label them as sepsis and led to use of higher empirical antibiotics. This fact will lead to undue use and overuse of antibiotics leading to increasing burden of antimicrobial resistance and creation of superbugs which will be extremely difficult to treat, creating an additional burden on already overburdened medical healthcare system, especially in a developing country.

NEWS-2 score was used to assess the organ dysfunction in this study which consists of different parameters related to patient without the need of any blood investigation or other laboratory parameters. NEWS-2 score was calculated for patients with mean score being 4.08 points [Table-11]. It was observed that mean NEWS-2 score was different with different categories of sepsis and different outcomes, with highest being for probable sepsis and patients who were discharged with unstable vitals. During data collection for study and also during analysis, it was observed that NEWS-2 score was sometimes falsely high in patients who had impaired level of consciousness due to non-infective causes (such as patients with dialysis dependent end stage renal disease, and decompensated chronic liver disease patients) and patients with chronic respiratory conditions having baseline low oxygen saturation and high baseline respiratory rate (such as patients with chronic obstructive pulmonary disease). It was also observed that the patients who required ICU admission had higher NEWS-2 score compared to its counterpart with mean NEWS-2 score being 7.09 and 3.75 respectively. These observations underscore the utility and limitations of the NEWS-2 score in sepsis identification. While it is effective for initial screening, certain patient conditions can lead to false positives, necessitating careful clinical judgment.

The overall mortality rate was 2.6% in the study participants which in absolute numbers was 6 mortalities, with equal distribution in both genders, i.e. 3 male and 3 female patients [Table-11]. This difference in outcomes of patients with suspected sepsis was majorly due to having patients admitted in ward, rather than an ICU facility, which would have been associated with a higher mortality rate.

### Limitations

Every research done is bound to have some or other limitations, and this one is no exception from this rule. Despite all measures, there are associated limitations with this study which highlighted below.

⍰ Single-Center Setting: This study was conducted at a single tertiary care center and conducting the study at a single center may limit the generalizability of the findings to a broader population. The results might reflect specific characteristics of the study site or patient population, reducing the external validity and potentially affecting the achievement of the research aims.
⍰ Use of newly prepared approach: The 3-step approach/ model prepared for this study was not previously utilised or studied in any study, making its application in real world scenario can be challenging.
⍰ Non-availability of reference study: Utilisation of a newly self-prepared approach was used in this study with no prior available reference study for efficacy of the approach adds to its limitations.
⍰ External validity: The study’s findings may not be applicable to populations with different demographic characteristics or healthcare practices, as it was conducted in a specific tertiary care center. Therefore, the external validity and generalizability of the results to other settings need to be carefully considered.

Limitations should not halt the process of research, rather it paves a way to overcome these limitations and make upcoming research more robust than the previous one.

### Interpretation

This study underscores the importance of a structured approach to sepsis identification and classification, especially in resource-limited settings. The 3-step model, combined with the NEWS-2 score, provides a practical framework for clinicians to identify sepsis early and accurately. With the use of this approach, we were able to classify patients into different sepsis categories, and found that on Day-1, the majority of patients were classified into probable sepsis (51.3%) and possible sepsis (35.2%), out of which majority of patients were later classified into asepsis group, when analysed over different time intervals of their hospital admission. However, the study also highlights the limitations of the NEWS-2 score, particularly in patients with chronic conditions or non-infective causes of impaired consciousness. This study also highlights the dynamic nature of sepsis, changing with duration and intervention (medical or surgical) and the increasing burden of antimicrobial resistance with overuse and irrational use of antibiotics, and to follow antibiotic stewardship.

Future research should focus on refining sepsis diagnostic criteria and developing more specific tools that can differentiate sepsis from other conditions with similar presentations. Additionally, incorporating molecular and biomarker-based approaches may enhance diagnostic accuracy. Ultimately, improving early sepsis identification and management can significantly reduce the associated morbidity and mortality, aligning with global health priorities set by the WHO.

Hence, sepsis identification and classification/categorization is a dynamic process. The present study could prove this dynamic process at admission, and every weekly interval till discharge/death outcomes. Despite it’s difficulties, this 3-step model holds true in identifying sepsis and in classifying them. Future multi-centric validation study will prove it’s exactness.

### Generalisability

As previous discussed in limitation section, the study’s findings may not be applicable to populations with different demographic characteristics or healthcare practices, as it was conducted in a specific tertiary care center. Therefore, the external validity and generalizability of the results to other settings need to be carefully considered, and can be commented with confidence only after study with larger sample size and including participants from different demographic background.

## Data Availability

All data produced in the present study are available upon reasonable request to the authors

